# Efficacy and safety of interferon β-1a in treatment of severe COVID-19: A randomized clinical trial

**DOI:** 10.1101/2020.05.28.20116467

**Authors:** Effat Davoudi-Monfared, Hamid Rahmani, Hossein Khalili, Mahboubeh Hajiabdolbaghi, Mohamadreza Salehi, Ladan Abbasian, Hossein Kazemzadeh, Mir Saeed Yekaninejad

**Affiliations:** Department of Pharmacotherapy, Imam Khomeini Hospital Complex, Tehran University of Medical Sciences, Tehran, Iran; Department of Infectious Diseases, Imam Khomeini Hospital Complex, Tehran University of Medical Sciences, Tehran, Iran; Advance Thoracic Research Center, Occupational Sleep Research Center, Tehran University of Medical Sciences, Tehran, Iran; Department of Epidemiology and Biostatistics, School of Public health, Tehran University of Medical Sciences, Tehran, Iran

**Author notes:** Corresponding Author Hossein Khalili, Professor of Pharmacotherapy, Department of Pharmacotherapy, Tehran University of Medical Sciences, Tehran, Iran. Postal Code: 1417614411 P.O.Box: 14155/6451.

**Keywords:** COVID-19, Interferon, Clinical response, Mortality

## Abstract

**Objectives:** To the best of our knowledge, there is no published study regarding use of IFN β-1a in the treatment of severe COVID-19. In this randomized clinical trial efficacy and safety of IFN β-1a has been evaluated in patients with severe COVID-19.

**Methods:** Forty-two patients in the interferon group received IFN β-1a in addition to the standard of care. Each 44 micrograms/ml (12 million IU/ml) of interferon β-1a was subcutaneously injected three times weekly for two consecutive weeks. The control group received only the standard of care. Primary outcome of study was time to reach clinical response. Secondary outcomes duration of hospital stay, length of ICU stay, 28-day mortality, effect of early or late administration of IFN on mortality, adverse effects and complications during the hospitalization.

**Results:** As primary outcome, time to the clinical response was not significantly different between the IFN and the control groups (9.7 ± 5.8 vs. 8.3 ± 4.9 days respectively, P = 0.95). On day 14, 66.7% vs. 43.6% of patients in the IFN group and the control group were discharged, respectively (OR = 2.5; 95% CI: 1.05-6.37). The 28-day overall mortality was significantly lower in the IFN then the control group (19% vs. 43.6% respectively, p = 0.015). Early administration significantly reduced mortality (OR = 13.5; 95% CI: 1.5–118).

**Conclusion:** Although did not change time to reach the clinical response, adding to the standard of care significantly increased discharge rate on day 14 and decreased 28-day mortality.

## Introduction

Severe acute respiratory syndrome –coronavirus-2 (SARS-CoV-2) is a novel virus that has been introduced as the first pandemic of the century, since its first detection in late December, 2019 in China [1]. The disease is named as corona virus disease 2019 (COVID-19) and manifests with dyspnea, fever, cough, myalgia and other flu-like symptoms [2]. However, it can progress to more severe disease and causes acute respiratory distress syndrome (ARDS), organ failure and death [3]. In the absence of definite treatment, the disease has caused more than 300,000 deaths worldwide in less than five months [4].

Some antivirals are examining in treatment of COVID-19. However, the results are not sound. Data in term of efficacy of lopinavir-ritonavir were promising at first but recent randomized trial failed to show benefits, especially in later stages of the disease [5]. Hydroxychloroquine is another available choice that has been included in many national recommendations and protocols [6]. However, it has also been advised to restrict this drug for clinical trials due to its questionable efficacy and risk of adverse effects [7]. The race is still on to find an effective treatment for COVID-19. Most of the experiences came from other corona virus epidemics; severe acute respiratory syndrome (SARS) and Middle East respiratory syndrome (MERS) [8].

Interferon (IFN) subtypes were previously examined in treatment of SARS and MERS. Primary in-vitro experience showed antiviral effects of IFNs, especially IFN-β and IFN-γ on SARS-CoV [9]. Also, same results were reported for IFN-β against MERS-CoV [10–11]. Later, in an animal model, higher antiviral activity of IFN-β γ compared to lopinavir-ritonavir was shown against MERS-CoV [12]. The efficacy of IFN-β on MERS is still being investigated [13].

The antiviral effect of IFNs express through activating interferon-stimulated gene (ISG) that slow viral replication [14]. Additionally, by decreasing the vascular leakage, IFN β-1a improved ARDS complications, regardless of its antiviral properties [15]. Also the higher expression of a protein named CD-73 could lead to better prognosis in ARDS. However, this data was not replicated in larger trial [16].

To the best of our knowledge, there is no published study regarding use of IFN β-1a in the treatment of severe COVID-19. In this randomized clinical trial efficacy and safety of IFN β-1a has been evaluated in patients with severe COVID-19.

## Methods

### Study design

This open-label randomized clinical trial was conducted to assess efficacy and safety of IFN-β-1a in treatment of adult (aged ≥ 18 years old) patients diagnosed with COVID-19. Patients were admitted during 29th February to 3^rd^ April 2020 in Imam Khomeini Hospital Complex, as main central hospital in Tehran, capital of Iran.

### Eligibility criteria

The diagnosis of COVID-19 was according to either a positive Real-Time Polymerase Chain Reaction (RT-PCR) of the respiratory tract samples or clinical signs/symptoms and imaging findings highly suspicious for COVID-19. Patients with the severe disease according the previous definition [17] with following criteria were included: (1) hypoxemia (need for noninvasive or invasive respiratory support to provide capillary oxygen saturation above 90%) (2) Hypotension (systolic blood pressure less than 90 mmHg or vasopressor requirement) (3) renal failure secondary to COVID-19 (according to KDIGO definition) [18] (4) neurologic disorder secondary to COVID-19 (decrease of 2 or more scores in Glasgow Coma Scale) (5) thrombocytopenia secondary to COVID-19 (platelet count less than 150000 /mm3) (6) severe gastrointestinal symptoms secondary to COVID-19 (vomiting/diarrhea that caused at least mild dehydration). Patients with each of the following characteristics were excluded: allergy to IFNs, receiving IFNs for any other reasons, previous suicide attempts, alanine amino transferase (ALT)> 5× the upper limit of the normal range and pregnant women.

Ethics Committee of Tehran University of Medical Sciences approved the study (approved ID: IR.TUMS.VCR.REC.1398.1052). Also the protocol of trial was registered (Registered ID: IRCT20100228003449N28).

The protocol of study was explained for all patients or their caregivers and informed consent was obtained from each participant. Eligible patients were allocated to IFN or control group using block randomization method.

### Treatment protocols

Patients in the IFN group received IFN β-1a in addition to the standard of care. Each 44 micrograms/ml (12 million IU/ml) of interferon β-1a (ReciGen®, CinnaGen Co., Iran) was subcutaneously injected three times weekly for two consecutive weeks. The control group received only the standard of care. The standard of care (the hospital protocol) consisted of hydroxychloroquine (400 mg BD in first day and then 200 mg BD) plus lopinavir/ritonavir (400/100 mg BD) or atazanavir/ritonavir (300/100 mg daily) for 7–10 days. Also primary care, respiratory support, fluid, electrolytes, analgesic, antipyretic, corticosteroid and antibiotic were recommended in the hospital protocol if indicated. The duration of study was two weeks and the patients were followed up to four weeks.

Demographic data, baseline characteristics and laboratory data were recorded for each patient. APACHE II score at the time of ICU admission was calculated for critically ill patients. All patients were daily monitored in term of vital signs, medical interventions and clinical conditions during the study course.

Need for respiratory support (invasive, non-invasive or no required) were assessed by a physician, regularly. Patients were assessed to fit in one of the six-category ordinal scale at days 0, 7, 14 and 28 of the inclusion [19]. If discharged, patient was followed by phone. Readmission was surveyed until 3^rd^ May.

### Outcomes assessment

Primary outcome of study was time to reach clinical response. Clinical response was defined according to the six-category ordinal scale [19]. This scale classifies patients in six categories according to the severity of the viral pneumonia. The six categories are: (1) discharge (2) hospital admission, not requiring oxygen (3) hospital admission, requiring oxygen (4) hospital admission, requiring non-invasive positive pressure ventilation (5) hospital admission requiring invasive mechanical ventilation (6) death. Time to clinical response was considered days required to at least two scores improvement in the scale or patient’s discharge, which one that occurred sooner. Secondary outcomes were duration of mechanical ventilation, duration of hospital stay, length of ICU stay, 28-day mortality, effect of early or late (before or after 10 days of onset of the symptoms) administration of IFN on mortality, adverse effects and complications during the hospitalization. Following adverse effects of the antiviral regimen/IFN β-1a and complications during the hospitalization course were assessed: gastrointestinal (nausea, vomiting, diarrhea, abdominal pain, pancreatitis), anaphylaxis and allergic reactions (rash, urticaria, angioedema, bronchospasm, and dyspnea related to medication administration), IFN injection-related reaction (skin erythema and necrosis, chills, fever, and flu-like symptoms after injection), neuropsychiatric (sleep disorder, psychosis, agitation, depression, and mania), renal impairment (according to KDIGO definition) [17], hepatic impairment (hepatic aminotransferases serum levels raised more than three times the upper limit of normal or serum total bilirubin above 2 mg/dL) [20], Indirect hyperbilirubinemia (direct bilirubin level less than 15% of the total bilirubin) [21], incidence of thromboembolism (deep vein thrombosis or pulmonary thromboembolism), incidence of nosocomial infections, diagnosis of septic shock (according to surviving sepsis campaign guideline) [22]. The Naranjo scale was used for evaluation of adverse effects of IFN [23].

### Statistical analysis

The quantitative variables were reported as mean ± standard deviation (SD) or as median with Inter Quartile Range (IQR). The qualitative ones were reported as number (percentage). For comparing the quantitative variables, independent sample *t* test was used in the case of normal distribution. Otherwise, Mann-Whitney test was applied. The qualitative variables were compared by Chi-square test.

Sample size was estimated for medium effect size with α = 0.05 and power = 0.85. The randomization was performed by permuted block randomization and block sizes of 2, 4 and 6 used randomly.

Days to reach the clinical response were extracted by Kaplan-Meier plot and were compared with a log-rank test. The HR and 95% CI for clinical improvement and HR with 95% CI for clinical death were calculated by Cox proportional hazards model. The odds ratio was also calculated for patients who received IFN early vs. late (after 10 days of starting the symptoms). SPSS software (version 21.0) was used for statistical analyses.

## Results

### Patients and baseline features

During the study period, 139 patients were screened, of whom 103 subjects were eligible. Considering dropped-outs, finally 81 patients (42 in the IFN and 39 in the control group) completed the study (figure 1). Males were 54.3% of patients. The mean ± SD of age in the IFN and control groups was 56.0 ± 16 and 59.5 ± 14 years, respectively. Hypertension (38.3%), cardiovascular diseases (28.4%), diabetes mellitus (27.2%), endocrine disorders (14.8%), and malignancy (11.1%) were common baseline diseases. Endocrine disorders were dyslipidemia and hypothyroidism. There was no significant difference in terms of demographic data and baseline diseases between the groups. The most frequent chief complaints of patients were cough, fever and dyspnea (table 1). APACHE II score at the time of ICU admission was not significantly different between two groups (15.3 ± 4 in the IFN group vs. 14.5 ± 3 in the control group, p = 0.79)

**Table 1.** Baseline characteristics of patients.

| Variable | IFN group (42) | Control group (39) | p-value |
| --- | --- | --- | --- |
| Age (mean ± SD) (year) | 56.09 ± 16 | 59.53 ± 14 | 0.61 |
| Male | 22 (52.38) | 21 (53.84) | 0.56 |
| Female | 20 (47.61) | 17 (43.58) |  |
| BMI (kg/m<sup>2</sup>), (mean ± SD) | 26.57 ± 5 | 26.69 ± 3 | 0.75 |
| Baseline diseases: n (%) |  |  |  |
| Any comorbidity | 32 (76.19) | 31 (79.48) | 0.46 |
| Hypertension | 15(35.71) | 16(41.02) | 0.39 |
| Diabetes mellitus | 13(30.95) | 9(23.07) | 0.29 |
| Ischemic heart disease | 11(26.19) | 12(30.76) | 0.41 |
| Endocrine disorder | 6(14.28) | 6(15.38) | 0.56 |
| Malignancy | 4(9.52) | 5(12.82) | 0.45 |
| Neuropsychiatric disorders | 3(7.14) | 2(5.12) | 0.53 |
| Hematologic Disorder | 2 (4.76) | 0 | 0.26 |
| Rheumatoid disorder | 1 (2.38) | 2 (5.12) | 0.47 |
| Renal disease | 1(2.38) | 2(5.12) | 0.47 |
| Liver disease | 1(2.38) | 2 (5.12) | 0.47 |
| Rheumatoid Arthritis | 1(2.38) | 1(2.56) | 0.73 |
| Asthma | 1(2.38) | 0 | 0.51 |
| Transplantation | 1(2.38) | 0 | 0.51 |
| COPD | 0 | 1(2.56) | 0.48 |

| Symptoms at admission: n (%) |  |  |  |
| --- | --- | --- | --- |
| Cough | 37 (88.09) | 25(64.10) | 0.011 |
| Fever | 30(71.42) | 20(51.28) | 0.05 |
| Dyspnea | 29(69.04) | 27(69.23) | 0.58 |
| Myalgia | 16 (38.09) | 20(51.28) | 0.16 |
| Chills | 16 (38.09) | 5 (12.82) | 0.009 |
| Anorexia | 12(28.57) | 6 (15.38) | 0.12 |
| Diarrhea | 8 (19.04) | 2(5.12) | 0.05 |
| Malaise | 6 (14.28) | 8 (20.51) | 0.32 |
| Nausea/Vomiting | 4(9.52) | 10(25.64) | 0.05 |
| Headache | 3 (7.14) | 4 (10.25) | 0.45 |
| Chest discomfort | 1 (2.38) | 4 (10.25) | 0.15 |
| Duration of symptoms before admission (mean ± SD) | 8.33 ± 4.5 | 6.57 ± 3.6 | 0.41 |

BMI: Body Mass Index, COPD: Chronic Obstructive Pulmonary Disease, SD: Standard Deviation

**Figure 1:**
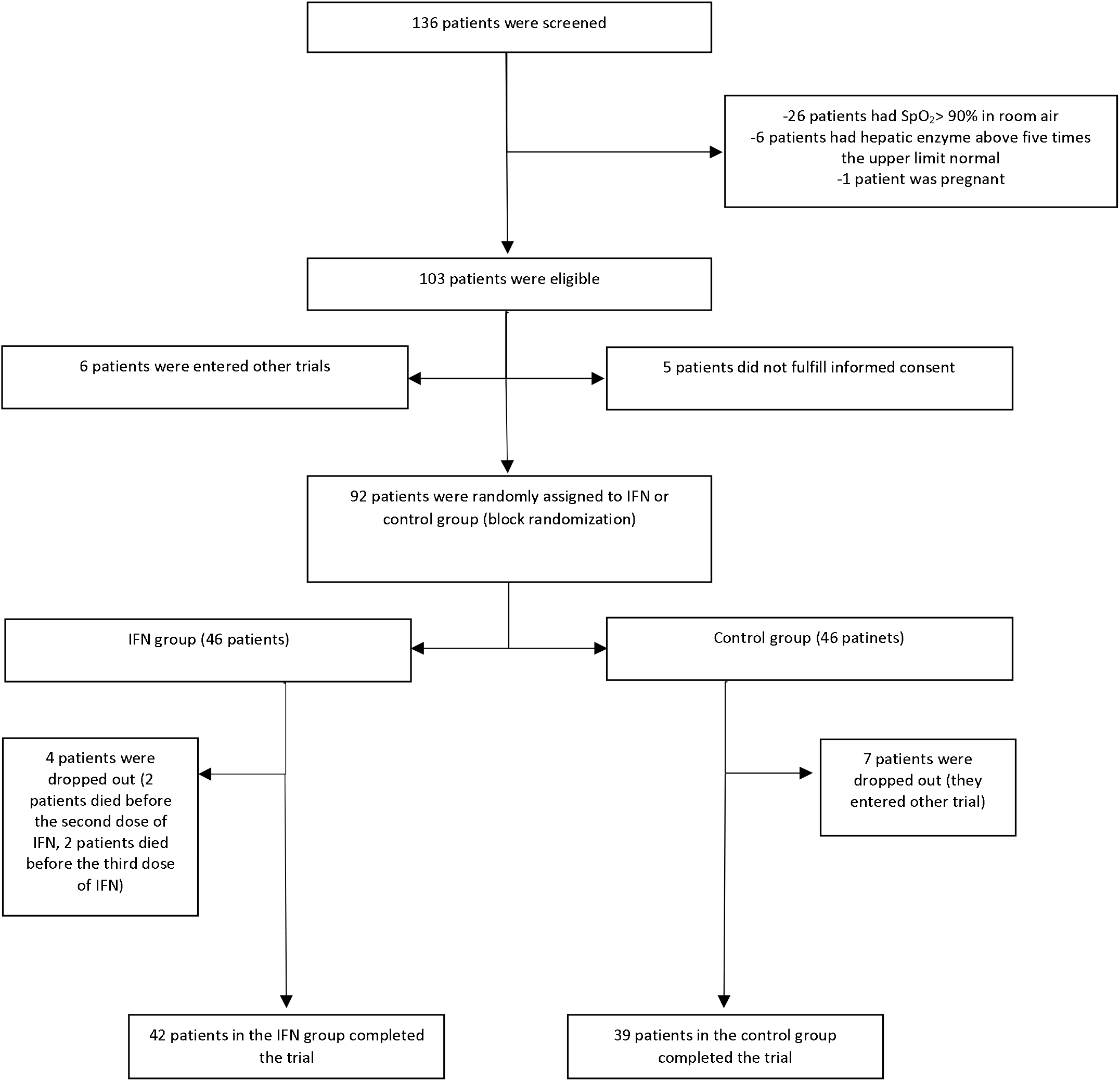
Consort flowchart of the study

### Vital signs and laboratory data

Median time from starting the symptoms (according to patients’ reports) to start IFN was 10 days (IQR: 8–13). The vital signs at the time of hospital admission were not statistically different, except respiratory rate that was significantly higher in the IFN group (24 vs. 22 respectively, p = 0.009). Comparing the baseline laboratory data at the time of hospital admission revealed that mean ± SD of blood urea nitrogen level was higher in the INF than the control group (36 ± 21 vs. 17 ± 8 respectively, p<0.001). Other laboratory findings were comparable (table 2)

**Table 2.** Initial vital signs and laboratory data Variable IFN group (Mean ± SD)

| Variable | IFN group (Mean ± SD) | Control group (Mean ± SD) | p-value |
| --- | --- | --- | --- |
| Temperature ($^{\circ}$C) | 37.50 ± 1.07 | 37.49 ± 0.98 | 0.48 |
| Heart rate (beats /minute) | 93 ± 15 | 90 ± 15 | 0.96 |
| Respiratory rate (breaths /minute) | 24 ± 6 | 22 ± 4 | 0.009 |
| Systolic blood pressure (mm Hg) | 122 ± 15 | 120 ± 14 | 0.66 |
| SPO<sub>2</sub> (%) | 86 ± 6 | 85 ± 7 | 0.86 |
| White Blood Cell (cells /μl) | 8345± 4632 | 7686 ± 4033 | 0.62 |
| Acute Lymphocyte count (cells /μl) | 1058 ± 449 | 935 ± 535 | 0.91 |
| Hemoglobin(g/dl) | 12.77 ± 2.23 | 13.02 ± 2.06 | 0.56 |
| Platelet count (cells $\times 10^3$/ μl) | 210 ± 88 | 200 ± 67 | 0.07 |
| Blood Urea Nitrogen (mg/dl) | 36 ± 21 | 17 ± 8 | 0.000 |
| Creatinine(mg/dl) | 1.15 ± 0.33 | 1.10 ± 0.32 | 0.80 |
| Sodium(meq/l) | 138 ± 3 | 137 ± 4 | 0.96 |
| Potassium(meq/l) | 4.29 ± 0.63 | 4.20 ± 0.58 | 0.47 |
| Calcium(mg/dl) | 8.12 ± 0.68 | 8.17 ± 0.49 | 0.18 |
| Phosphorus(mg/dl) | 3.30 ± 0.93 | 2.83 ± 0.65 | 0.06 |
| Magnesium(mg/dl) | 2.08 ± 0.36 | 1.96 ± 0.28 | 0.25 |
| Aspartate aminotransferase(u/l) | 50 ± 32 | 50 ± 34 | 0.88 |
| Alanine aminotransferase(u/l) | 44 ± 37 | 41 ± 26 | 0.66 |
| Alkaline phosphatase(u/l) | 184 ± 96 | 203 ± 168 | 0.36 |
| Total bilirubin(mg/dl) | 0.81 ± 0.54 | 1.14 ± 1.42 | 0.22 |
| International Normalized Ratio<br>(INR) | 1.09 ± 0.11 | 1.55 ± 1.44 | 0.000 |
| C-reactive protein(mg/dl) | 139 ± 74 | 121 ± 78 | 0.56 |
| Erythrocyte sedimentation<br>rate(mm/h) | 77 ± 32 | 63 ± 28 | 0.96 |
| Lactate dehydrogenase(u/l) | 781 ± 412 | 809 ± 379 | 0.49 |
| Creatine phosphokinase(u/l) | 288 ± 339 | 316 ± 698 | 0.60 |
| Troponin-I(ng/l) | 11 ± 17 | 46 ± 175 | 0.17 |

### Treatment strategies

Hydroxychloroquine is main medication in Iran’s national protocol for the treatment of COVID-19. lopinavir/ritonavir or atazanavir/ ritonavir may be added to hydroxychloroquine in severe cases. The antiviral regimens were not significantly different between two groups. Deep vein thrombosis prophylaxis and stress ulcer prophylaxis were considered for patients if indicated. Based on patients’ clinical conditions, azithromycin, intravenous ascorbic acid, antibiotics, intravenous immunoglobulin or a corticosteroid were added to the antiviral regimens. A corticosteroid (methyl prednisolone, hydrocortisone or dexamethasone) was administered for 61.9% and 43.6% of patients in the IFN and the control groups, respectively. Also 35.7% and 25.6% of patients in the IFN and the control group received intravenous immunoglobulin respectively. Supportive care modalities and administered medications are summarized in table 3.

**Table 3.** Supportive care interventions and medications.

| Variable | IFN group (42) | Control group (39) | p-value |
| --- | --- | --- | --- |
| Time from starting the symptoms to start of the interventions (Mean ± SD) | 11.70 ± 5.71 | 9.31 ± 4.45 | 0.45 |
| ICU admission (%) | 19 (45.23) | 23 (58.97) | 0.25 |
| Respiratory support: n (%) |  |  |  |
| Nasal cannula | 2 (4.76) | 1 (2.56) | 0.65 |
| Face mask | 24 (57.14) | 21 (53.84) |  |
| NIPPV | 1 (2.38) | 0 |  |
| IMV | 15 (35.71) | 17 (43.58) |  |
| Medications: n (%) |  |  |  |
| Hydroxychloroquine | 40 (95.23) | 39(100.0) | 0.26 |
| Antiviral regimen | 42 (100) | 39 (100) | - |
| Azithromycin | 8 (19.04) | 5 (12.82) | 0.32 |
| Vitamin C | 13 (30.95) | 12 (30.76) | 0.58 |
| Broad spectrum antibiotics | 33 (78.57) | 27 (69.23) | 0.24 |
| Diphenhydramine | 17 (40.47) | 26 (66.66) | 0.016 |
| Antiemetic | 11 (26.19) | 6 (15.38) | 0.17 |
| Opioid | 14 (33.33) | 17 (43.58) | 0.23 |
| Stress ulcer prophylaxis | 42 (100) | 39 (100) | 0.54 |
| Deep vein thrombosis prophylaxis | 41 (97.61) | 37 (94.87) | 0.47 |
| Statins | 9 (21.42) | 6(15.38) | 0.34 |
| ARBs | 10 (23.80) | 4(10.25) | 0.09 |
| Beta-blockers | 8 (19.04) | 2 (5.12) | 0.06 |
| Calcium channel blockers | 7 (16.66) | 5 (12.82) | 0.43 |
| ACEIs | 1 (2.38) | 2(5.12) | 0.47 |
| Corticosteroid | 26 (61.90) | 17 (43.58) | 0.07 |
| Immunoglobulin | 15 (35.71) | 10 (25.64) | 0.23 |

ACEI: Angiotensin Converting Enzyme Inhibitor, ARB: Angiotensin Receptor Blocker, IMV: Invasive Mechanical Ventilation, NIPPV: Non-Invasive Positive Pressure Ventilation, SD: Standard Deviation

### Outcomes and complications

As primary outcome, time to the clinical response was not significantly different between the IFN and the control groups (9.7 ± 5.8 vs. 8.3 ± 4.9 days respectively, P = 0.95). It has been shown in the Kaplan-Meier plot (figure 2). Also the log rank test revealed that there was no statistically significant difference between the groups, considering time to clinical response (HR = 1.10; 95% CI: 0.64–1.87, p = 0.72).

**Figure 2:**
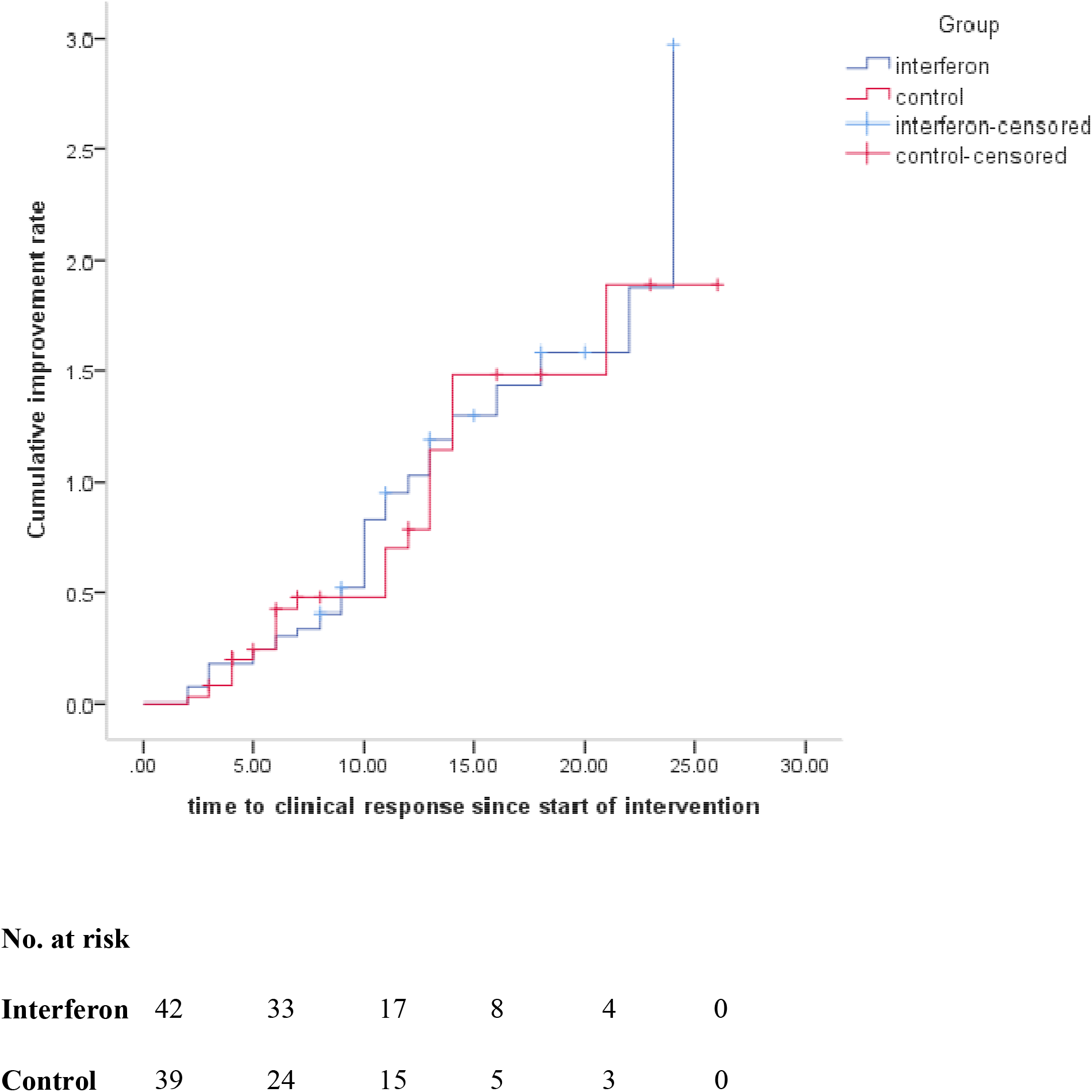
Time to clinical response. Hazard Ratio was calculated as 1.10 with 95% CI 0.64-1.87.

The six-category ordinal scale was assessed at days 0, 7, 14 and 28 (table 4). On day 0, there was no significant difference between the groups in terms of the components of this scale. On day 7 of therapy, 19% of patients in the INF group were discharged and with no death. At this time, 28% of patients in the control group were discharged and 25% died. However the difference was not statistically considerable (OR = 0.60; 95% CI: 0.21-1.69). On day 14, the results were statistically notable and 66.7% vs. 43.6% of patients in the IFN group and the control group were discharged, respectively (OR = 2.5; 95% CI: 1.05-6.37).

**Table 4.**
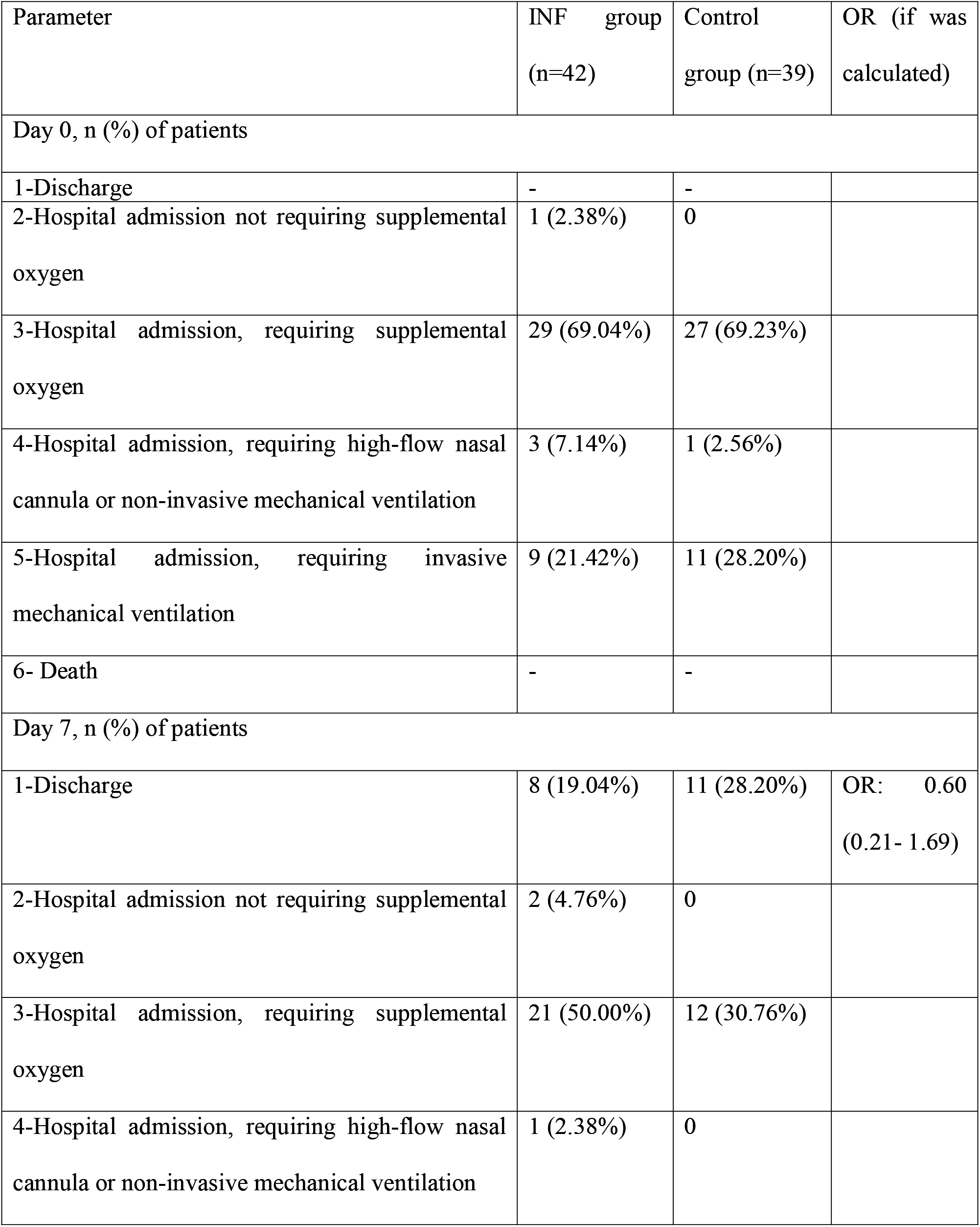
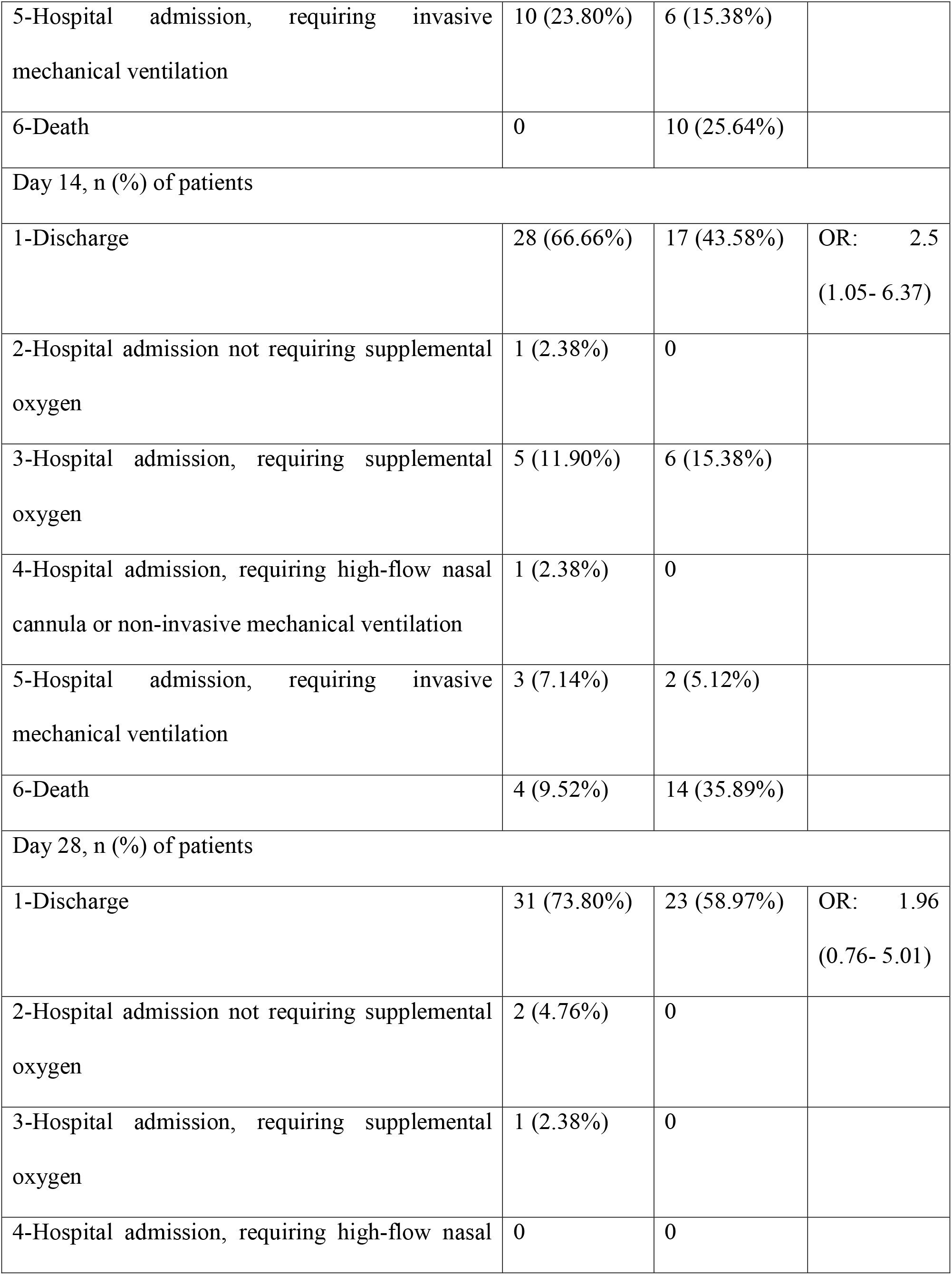
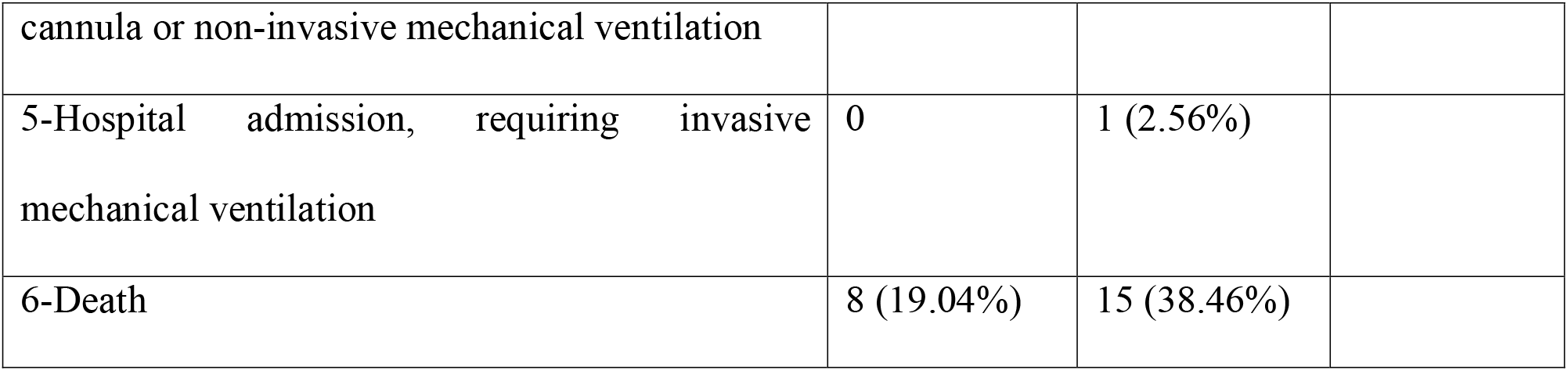
Findings based on the six-category scale at days 0, 7, 14, 28.

Regarding the time of INF initiation, the analysis showed that early administration significantly reduced mortality (OR = 13.5; 95% CI: 1.5–118). However, late administration of INF did not show significant effect (OR = 2.1; 95% CI: 0.48–9.6).

Other secondary outcomes such as duration of hospital stay, length of ICU stay and duration of mechanical ventilation were not statistically different. However, more patients were extubated in the IFN group (p = 0.019). Additionally, the 28-day overall mortality was significantly lower in the IFN then the control group (19% vs. 43.6% respectively, p = 0.015).

Complications during the hospitalizations course, incidence of organ failure and adverse effects were not different between the groups. Injection-related side effects (fever, chills, myalgia and headache in few hours after injection of IFN) happened in 8 (19%) patients (table 5).

**Table 5.**
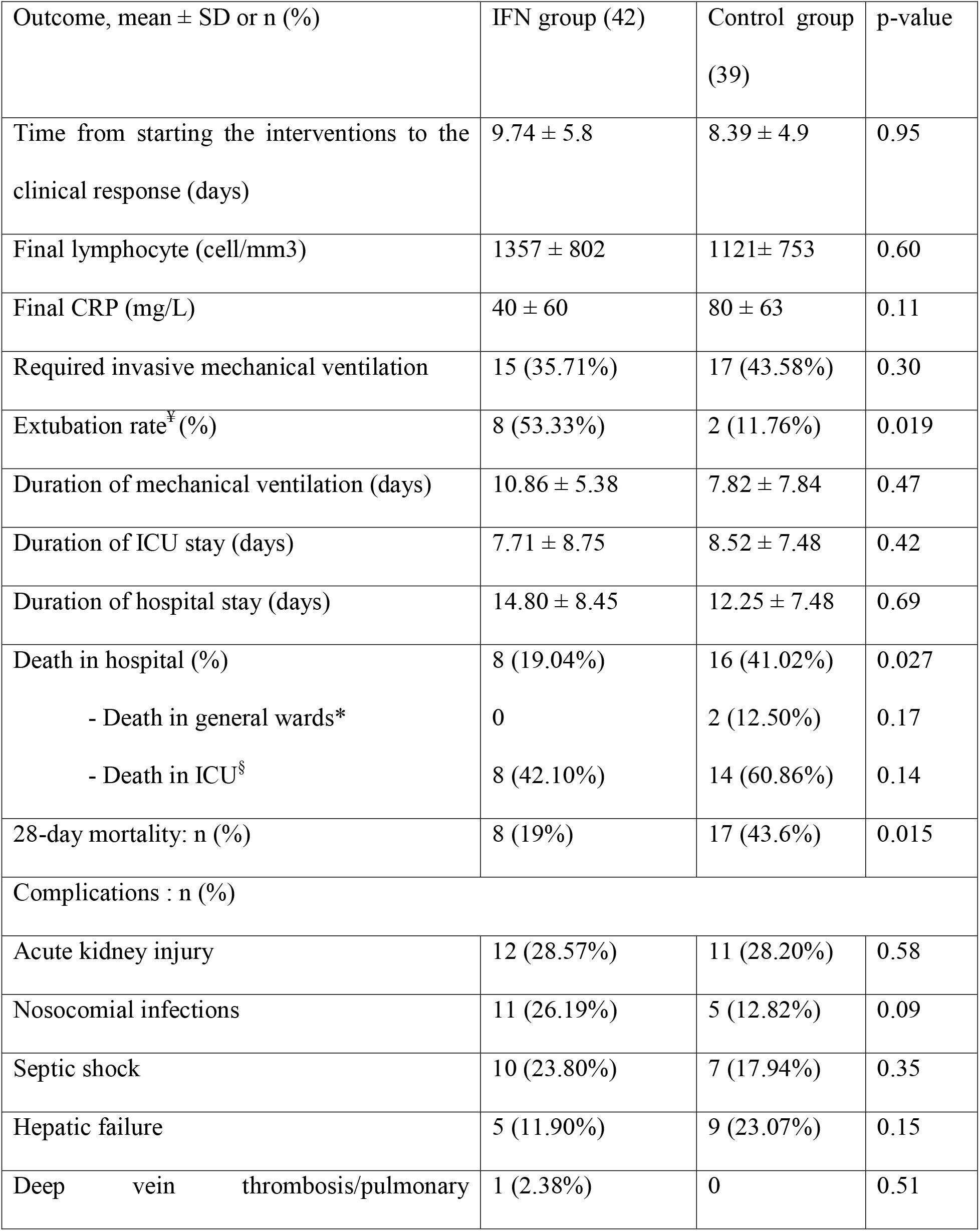
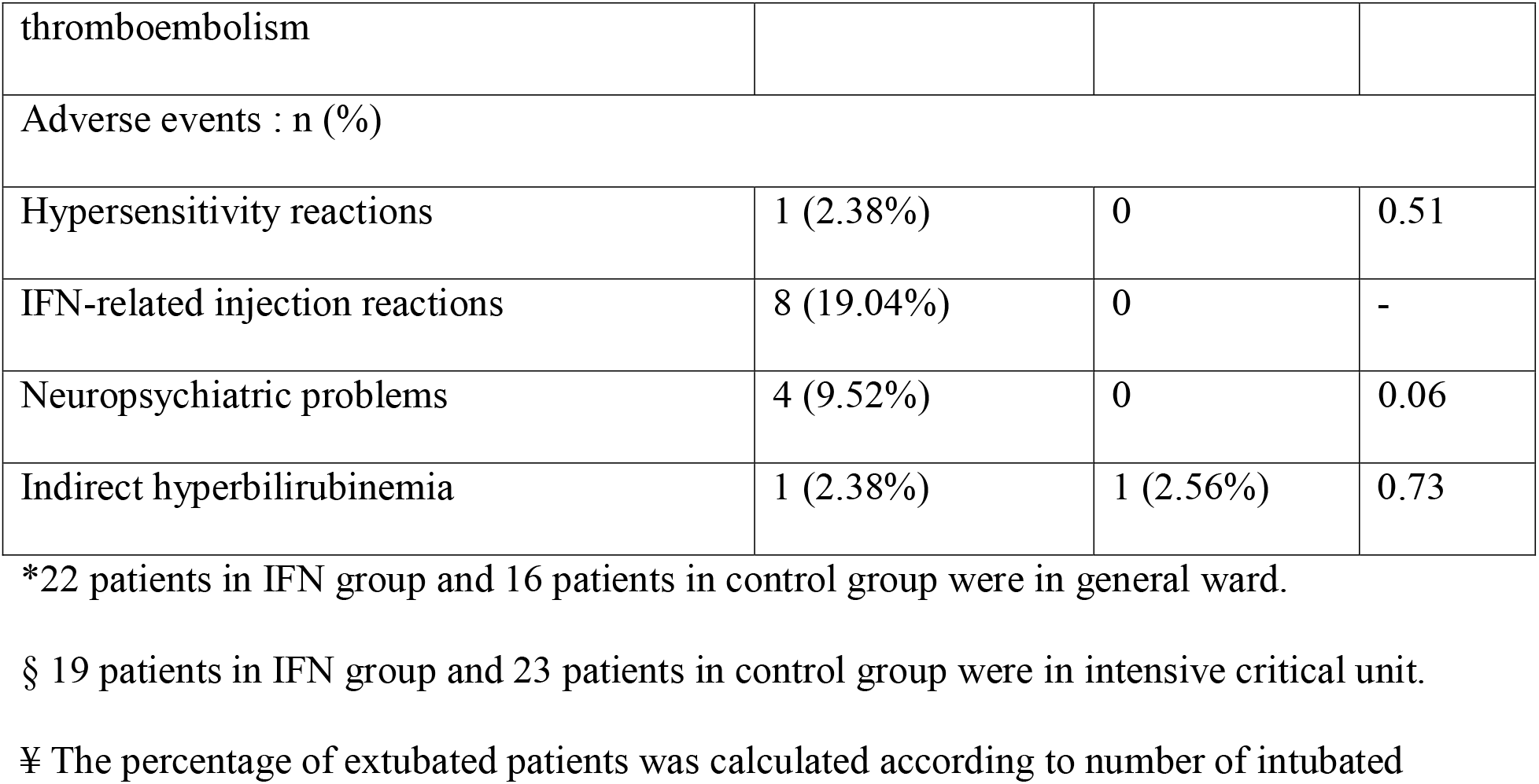
Outcomes of the study.

Hypersensitivity reaction was occurred in one patient who received IFN. The reaction was presented with maculopapular rash on trunk and both upper and lower limbs. Although the patient was taking herbal medicine for cough consisting of thyme and honey, and other medications like hydroxychloroquine and lopinavir-ritonavir concomitantly, interferon was discontinued after fourth dose and rashes begun to disappear within three days. According the Naranjo score the reaction was possibly due to IFN.

Neuropsychiatric problems were detected in 4 patients in the IFN group. Two cases experienced severe agitation and two cases complained of mood swing (mostly depression). One of the patients with mood swing had history of mild depressive disorder in past years. Out of 4 cases, two patients were being in the hospital for near to 1 month. Neuropsychiatric side effects of IFN are unlikely to happen in short-term use. All patients received psychiatry consult. According the Naranjo score, IFN possibly and probably caused neuropsychiatric problems in three and one patients, respectively.

## Discussion

Present study was first randomized open-label controlled trial that assessed the efficacy and safety of IFN β-1a in treatment of patients with diagnosis of severe COVID-19. Time to reach the clinical response did not change following adding IFN to the standard of care. However, IFN significantly improved discharge rate on day 14. Also 28-day mortality was significantly lower in the IFN group. Patients who received IFN in early phase of the disease experienced significantly more benefits of the treatment. Some injection-related adverse effects of IFN occurred and all were tolerable.

Still no effective therapeutic option has been introduced for COVID-19. Some anti-inflammatory agents and cytokine release inhibitors like corticosteroids and tocilizumab (acting against IL-6) have been proposed [24–25]. However increasing risk of secondary infections, activation of latent tuberculosis and other adverse effects are the serious concerns [26]. lopinavir-ritonavir did not improve time to the clinical improvement and mortality [5]. Efficacy of other therapeutic modalities like convalescent plasma is not clear [27].

Among coronaviruses family, the efficacy of IFNs at first was reported in SARS [28]. After subsidence of the SARS epidemic, IFNs was again proposed for treatment of another coronavirus, MERS. However, different subtypes of IFNs (alpha and beta) in combination with ribavirin did not show significant efficacy in critically ill patients with MERS [29]. Due to promising primary effects of IFN β-1 in MERS, a trial for evaluating the efficacy is still running [13]. Thus, IFNs, especially type I are still interesting options for recent epidemics. One study evaluated the effect of IFN β-1b in combination with lopinavir-ritonavir and ribavirin in mild to moderate COVID-19 [30]. Also, nebulized IFN α-2b in combination with oral arbidol was examined in treatment of the disease [31].

IFNs are natural cytokines that are produced in response to viral infections. They activate natural killer cells (NK) and macrophages. Interestingly, SARS-CoV showed ability to act against the effects of IFNs [32–33]. SARS-CoV encoded synthesis a family of proteins, Open Reading Frame (ORF) that inhibit STAT1 transporter to enter the nucleus and block the interferon signaling [33]. However, recently it has been shown that function of some proteins in this family (ORF6 and ORF3b) had changed in SARS-CoV-2. This may has changed pathogenesis of SARS-CoV-2 and its interaction with IFN [34].

Beside the antiviral effects of IFNs, potential role of IFN β-1a in improving ARDS complications was proposed. Expression of CD-73 proteins in lung cells and decrease in vascular leakage in ARDS and subsequent mortality was reported following treatment with intravenous IFN β-1a [15]. However, the results were not repeated in the next larger trial [16]. This may be related to extensive use of glucocorticoids in the latter trial that can interfere with effect of IFN. The antagonist effect of corticosteroids is considerable [35]. In our trial, some patients also received corticosteroids, but were not concomitant with IFN. Meanwhile corticosteroids were considered after the intervention, if required.

The mean age of patients in present study was 57± 15 years. The mean or median of age values were different in published studies from 46 to 65 years [36–37]. The male gender was dominant, resembling other studies in COVID-19 [5, 37]. Gender difference was evident in many studies of COVID-19. Mostly not affected outcomes, however, in a study, critically ill males had higher mortality [38]. This difference was first explained by increase in expression of ACE2 (the receptor for entrance of SARS-CoV2 to cell) in Asian men [39]. Later, the issue was contributed to higher rate of cigarette smoking in Asian men compared to Asian women. However, both hypotheses should be confirmed in future studies. At present neither gender nor smoking is certainly correlate to severity of COVID-19 [40].

Baseline vital signs and laboratory data were almost comparable between the groups, with some exceptions. The respiratory rate was significantly higher in the IFN group. Also, mean of blood urea nitrogen level was higher in this group. This may be due to higher rate of diarrhea as initial symptoms of COVID-19 in this group in comparison to the control one (19% vs 5.1%). Diarrhea may cause dehydration and lead to higher blood urea nitrogen.

As primary outcome of the study, time to reach the clinical response was not significantly different between the groups. In the study of Hung et al, clinical improvement significantly occurred faster in IFN combination therapy group (lopinavir-ritonavir + interferon β-1b + ribavirin) compared with the control group i.e. 7 vs. 12 days [5]. However, it should be noticed that in this study severe cases were excluded.

Length of ICU and hospital stay and duration of mechanical ventilation were not statistically different between the groups like the other interferon trial [30]. However, according to the six-category ordinal scale, more patients were discharged following IFN therapy at day 14. This scale was also used in the study of remdesivir, but same results were nor detected [37]. One of the remarkable findings in our study was decrease in 28-day mortality in IFN group that was not achieved in other studies in COVID-19 [30, 37].

The clinical course of COVID-19 is divided into early viral-replication phase and late cytokine-release phase [41]. It was suggested that early administration of antiviral medications (within 7-10 days of starting the symptoms) might improve outcome of patients with COVID-19[37]. Additionally, early administration of IFNs was recommended in treatment of MERS [42]. Early administration of antiviral agents in the viral infections can accelerate viral clearance and postpone neutrophil infiltration. Early administration of IFN β-1a, even in severely ill mechanically ventilated patients led to higher survival rate. Late administration did not show more benefits.

Regarding safety of IFN therapy in patients with COVID-19, incidence of injection-related reactions including fever, chills, headache and fatigue (early after injection) detected in 19% of the patients. All of these symptoms responded to the symptomatic therapy (acetaminophen) and change the time of injection to late night. Erythema or injection site reaction or any reaction that caused treatment interruption was not reported. Considering duration of the intervention, incidence rates of the adverse reactions were lower than that were reported in patients with multiple sclerosis [43]. However, it should be accounted that some patients in our study were under mechanical ventilation and exact evaluation of these reactions was not feasible. As a component of the supportive care in COVID-19, most patients received analgesic and antipyretic concomitant with antiviral agents and IFN. These medications might mask the adverse reactions of IFN, too.

Nausea, vomiting and abdominal pain were the most common gastrointestinal complications in our patients and the incidence rates were not different between the groups. Although two cases experienced slight elevation in serum amylase and lipase levels, in further evaluation no pancreatitis was confirmed. COVID-19 can cause several gastrointestinal symptoms. However, gastrointestinal symptoms that started after the hospital admission may be related to the medications. The incidence rates of AKI and hepatic impairment were not significantly different between the IFN and the control groups. Both renal and liver injuries can be COVID-19 associated organ dysfunction [44]. Also nephrotoxicity of medications like antibiotics and furosemide (that were frequently prescribed in our patients) and hepatotoxicity of antiviral agents should be taken into account. No case of hepatotoxicity that led to discontinuation of interferon was detected. Indirect hyperbilirubinemia is one of adverse effects of atazanavir-ritonavir [45].

This study had some limitations. Both patients in general, intermediate and intensive care units wards were recruited. Most of the general wards in fact were intermediate wards, but the accurate classification was not possible due to special and emergent conditions. Due to restrictions in each pandemic event and low experiences, diagnosis of COVID-19 was according to either positive RT-PCR or signs/symptoms plus imaging findings highly suggestive for the disease. Also considering the follow-up imaging and RT-PCR were not feasible.

## Conclusion

Although did not change time to reach the clinical response, adding to the standard of care significantly increased discharge rate on day 14 and decreased 28-day mortality. Improved survival was significant when patients received IFN β-1a in the early phase of the disease. Adverse effects of IFN β-1a were injection-related, neuropsychiatric problems and hypersensitivity reaction that all were tolerable and resolved during the follow-up period.

## Data Availability

Data available per request.

## Acknowledgement

We would like to thank the nurses and other staffs of Imam Khomeini Hospital Complex for their kind supports.

## Funding

The authors did not receive any fund for this work. ReciGen was a generous gift from CinnaGen Co.

## Declaration of interest

There is no conflict of interest for authors to declare.

## Notes

### Competing Interest Statement

The authors have declared no competing interest.

### Clinical Trial

IRCT20100228003449N28

### Funding Statement

The authors have not received any fund about this work.

### Author Declarations

Ethics Committee of Tehran University of Medical Sciences approved the study (approved ID: IR.TUMS.VCR.REC.1398.1052).

